# GAUSS: A comprehensive R package for accurate estimation of linkage disequilibrium for variants, Gaussian imputation and TWAS analysis of cosmopolitan cohorts

**DOI:** 10.1101/2023.09.19.23295783

**Authors:** Donghyung Lee, Silviu-Alin Bacanu

## Abstract

**Motivation:** As the availability of larger and more ethnically diverse reference panels grows, there is an increase in demand for ancestry-informed imputation of genome-wide association studies (GWAS), and other downstream analyses, e.g., fine-mapping. Performing such analyses at the genotype level is computationally challenging and necessitates access to individual-level genotype and phenotype data. Summary-statistics-based tools, not requiring individual-level data, provide an efficient alternative that streamlines computational requirements and promotes open science by simplifying the re-analysis and downstream analysis of existing GWAS summary data. However, existing tools perform only disparate parts of needed analysis, have only command-line interfaces and are difficult to extend/link by applied researchers.

**Results:** To address these challenges, we present GAUSS — a comprehensive and user-friendly R package designed to facilitate the re-analysis/downstream analysis of GWAS summary statistics. GAUSS offers an integrated toolkit for a range of functionalities, including i) estimating ancestry proportion of study cohorts, ii) calculating ancestry-informed linkage disequilibrium, iii) imputing summary statistics of unobserved variants, iv) conducting transcriptome-wide association studies, and v) correcting for “Winner’s Curse” biases. Notably, GAUSS utilizes an expansive, multi-ethnic reference panel consisting of 32,953 genomes from 29 ethnic groups. This panel enhances the range and accuracy of imputable variants, including the ability to impute summary statistics of rarer variants. As a result, GAUSS elevates the quality and applicability of existing GWAS analyses without requiring access to subject-level genotypic and phenotypic information.

**Availability and implementation:** The GAUSS R package, complete with its source code, is readily accessible to the public via our GitHub repository at https://github.com/statsleelab/gauss. To further assist users, we provided illustrative use-case scenarios that are conveniently found at https://statsleelab.github.io/gauss/.

**Supplementary information:** Supplementary data are available at *Bioinformatics* online.

## 1 Introduction

Genome-wide association studies (GWAS) have revolutionized our understanding of the genetic underpinnings of complex human diseases and traits. However, gaining access to individual-level genotype and phenotype data from these studies is often challenging due to privacy or regulatory constraints. Fortunately, summary statistics from GWAS are widely available and have catalyzed the development of various summary statistics-based genome analysis tools. Popular tools in this category include those for imputing summary statistics for unobserved genetic variants (Lee, et al., 2013; Pasaniuc, et al., 2014), prioritizing causal variants (Schaid, et al., 2018), computing polygenic risk scores (Lewis and Vassos, 2020), and conducting transcriptome-wide association studies (Gamazon, et al., 2015; Lee, et al., 2015). The effectiveness of these tools often hinges on the accurate estimation of cohort-specific linkage disequilibrium (LD)—the correlation structure among genetic variants. Often, researchers rely on publicly available reference panels like the 1000 Genomes panel, which encompasses genomic data from just 2,504 individuals across 26 ethnic groups. This scale and diversity are significantly limited compared to GWASs, which often involve hundreds of thousands of individuals from diverse ethnic backgrounds. Moreover, many of these tools operated solely in Linux command-line environments, posing a steep learning curve that deters many applied researchers from incorporating these valuable resources into their work.

To overcome these limitations, we present GAUSS (**G**enome **A**nalysis **U**sing **S**ummary **S**tatistics), a comprehensive R package designed for the analysis of multi-ethnic GWAS summary statistics. GAUSS utilizes a large multi-ethnic reference panel consisting of 32,953 genomes (33KG) (Chatzinakos, et al., 2021), including 20,281 Europeans, 10,800 East Asians, 522 South Asians, 817 Africans, and 533 Native Americans (See Supplementary Text 1 and Table 1 for further details). GAUSS equips researchers with a suite of powerful functionalities,

### Lee and Bacanu

including i) estimating the ethnic composition of a multi-ethnic GWAS cohort (Chatzinakos, et al., 2021; Lee, et al., 2015), ii) computing ancestry-informed linkage disequilibrium (LD), iii) imputing association Z-scores for unmeasured genetic variants (Lee, et al., 2013; Lee, et al., 2015), iv) performing transcriptome-wide association studies (Lee, et al., 2015; Lee, et al., 2016), and v) correcting for the “Winner’s Curse” (Bigdeli, et al., 2016) (See Figure 1A).

**Figure 1.**
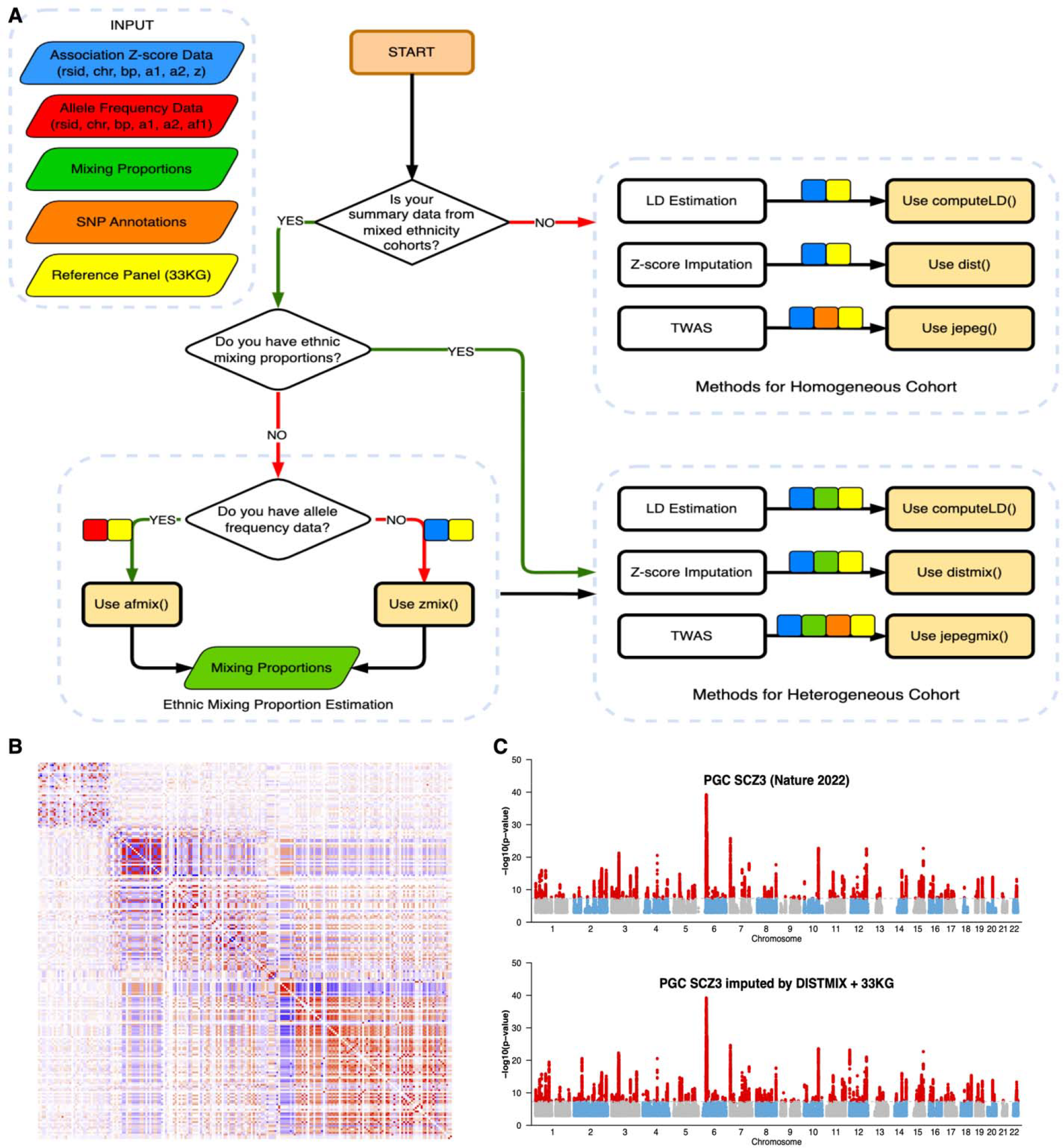
**A.** Flowchart Depicting GAUSS Analytical Procedure. GAUSS functions utilize five core input datasets: i) Association Z-score Data (highlighted in blue), consisting of six columns of variables: SNP ID (rsid), chromosome number (chr), base pair position (bp), reference allele (a1), alternative allele (a2), and Z-score (z); ii) Allele Frequency Data (colored in red), encapsulating rsid, chr, bp, a1, a2, and reference allele frequency (af1); iii) Ethnic Mixing Proportion Data (colored in green), comprising population name abbreviations and corresponding mixing proportions; iv) SNP Annotation Data (colored in orange), offering functional annotation information of SNPs; v) Reference Panel Data (colored in yellow), offering genotype information for a significant pool of 32,953 individuals from 29 diverse ethnic groups. Small colored squares on arrows pointing to GAUSS functions denote the required input data sets for each function. **B**. Ancestry informed LD of the 2022 PGC Schizophrenia GWAS cohorts (chromosome 10: 104-105Mb) **C**. Re-imputing PGC SCZ3 GWAS summary statistics using the ‘distmix()’ function, Illumina 1M SNP set and 33KG as a reference panel.

## 2. GAUSS R Package

### 2.1 Estimating ancestry proportions in GWAS cohorts

GAUSS enables users to estimate the ancestry proportions of the genetic association studies by utilizing allele frequencies (AF) through its ‘afmix()’ function (Lee, et al., 2015). In cases when allele frequencies for the cohort are not available, ‘zmix()’ can be used. This function is designed to estimate the ethnic composition using the association Z-scores (Chatzinakos, et al., 2021). These estimated ancestry proportions are particularly valuable for downstream analyses for multi-ethnic GWAS. (for more details on ‘afmix()’ function, refer to https://statsleelab.github.io/gauss/articles/afmix_example.html)

### 2.2. Estimating ancestry-informed LD

Genetic studies often include multi-ethnic samples to enhance detection power and resolution. Accurate estimation of ancestry-informed LD is essential in many tools using summary statistics from such diverse cohorts. To address this need, GAUSS offers the ‘computeLD()’ function, which calculates ancestry-adjusted LD values using our extensive 33KG reference panel. The function generates an LD matrix by calculating a weighted sum of LD values specific to each ethnic group. The LD matrix can be easily integrated into existing summary statistics-based analytical tools that require cohort-specific LD values. The ‘computeLD()’ function requires multiple inputs, such as chromosome number, start and end base pair positions for the window of interest, ancestry proportions, filenames for input and reference panels. Should users not have ancestry proportions, these can be approximated using the ‘afmix()’ and zmix() functions, as detailed in the previous section. For further guidance on the ‘computeLD()’ function, refer to: https://statsleelab.github.io/gauss/articles/computeLD_example.html

### 2.3. Imputing summary statistics of unmeasured SNPs

As the scale and ethnic diversity of reference panels continue to expand, the need for imputing/re-imputing data becomes increasingly critical. GAUSS addresses these challenges by offering two specialized functions, ‘dist()’ and ‘distmix()’, designed for directly imputing summary statistics (i.e., association Z-scores) of unmeasured SNPs in both homogeneous and multi-ethnic cohorts (Lee, et al., 2013; Lee, et al., 2015). Utilizing the 33KG reference panel, these functions facilitate highly accurate imputations. The ‘distmix()’ function is particularly optimized to handle cohorts with varying ethnic compositions. The imputed summary statistics are readily usable in a wide range of downstream analyses, including transcriptome-wide association studies and meta-analyses. For further details, refer to https://statsleelab.github.io/gauss/articles/dist_example.html.

### 2.4. Transcriptome-wide association studies

Transcriptome-Wide Association Studies (TWAS) serves as a powerful approach to explore the functional links between genetic variations and complex traits by synergizing GWAS findings with functional annotations. GAUSS facilitates these analyses by incorporating advanced TWAS tools, ‘jepeg()’ and ‘jepegmix()’, designed for both homogeneous and heterogeneous cohorts, respectively (Lee, et al., 2015; Lee, et al., 2016). The ‘jepeg()’ function performs TWAS by testing for the joint effect on phenotype of expression quantitative trait loci (eQTLs) and functional variants in a gene, leveraging the 33KG reference panel. It requires SNP annotation data that provides essential mapping between SNPs and their corresponding gene-level functional information. It integrates information from the SNP annotation file to construct gene-level test statistics, aiming to identify genes whose expression levels are potentially modulated by genetic variations. The ‘jepegmix()’ function extends the capabilities of ‘jepeg()’ to multi-ethnic cohorts. The results generated by these functions include gene-wise association p-values, estimated effect sizes, and other relevant statistics, offering valuable resources for subsequent functional validation or integrative analyses. For more details, refer to https://statsleelab.github.io/gauss/articles/jepeg_example.html.

### 2.5. Winner’s curse adjustment

In genetic research, ‘sub-threshold’ association signals often have a larger impact on trait variance than statistically significant variants. Accurately estimating these effects is crucial, yet challenging due to “winner’s curse” biases. Previously, we’ve introduced a simple adjustment method called FIQT (**F**DR **I**nverse **Q**uantile **T**ransformation), to adjust for these biases using only Z-scores, which represent the true means or noncentralities (Bigdeli, et al., 2016). FIQT is now conveniently integrated as the ‘fiqt()’ function in the GAUSS package. By simply inputting a Z-score vector and the minimum non-zero p-value output by qnorm() function (10^-320^ by default), the function provides a Z-score vector adjusted for the winner’s curse. For more details, refer to https://statsleelab.github.io/gauss/articles/fiqt_example.html.

### 2.5 Applying GAUSS to the 2022 PGC Schizophrenia GWAS

To demonstrate the practical utility of GAUSS, we applied its functions to the 2022 Psychiatric Genomics Consortium (PGC) Schizophrenia (SCZ) GWAS dataset (Trubetskoy, et al., 2022).

To showcase the applicability of ‘computeLD()’ function, we first computed ancestry-informed LD using the 33KG panel and PGC SCZ 2022 data for a genomic region chr10:104-105Mb. The genetic correlation between single nucleotide polymorphisms (SNPs) within this region is visually represented via an intuitive heatmap (Figure 1B).

Next, we utilized GAUSS’s distmix() function to re-impute summary statistics for unmeasured SNPs in the dataset. We selected Illumina 1M SNPs as our baseline data for measured SNPs. The afmix() function was first applied to estimate the ethnic composition of the study cohort based on this 1M SNP dataset. These estimated proportions, along with the association summary statistics of the measured SNPs, were then used as inputs for the distmix() function to generate association Z-scores for SNPs absent in the Illumina 1M array. We run the imputation for 2,724 1Mega base pair regions using the Miami University Linux Cluster. The Manhattan plot (Figure 1C) is created based on the results and compared to the results from the original study. As shown in the figure, distmix() successfully re-imputed the 2022 PGC SCZ GWAS data using the 33KG panel and identified more LD independent association regions not found from the original study.

## 3. Conclusion

We have developed the R package GAUSS designed for the re-analysis/downstream analysis of summary statistics from large-scale multi-ethnic genetic association studies. Unlike traditional tools that often rely on Linux command-line interfaces, GAUSS provides a user-friendly experience within the widely used R environment. Built upon robust R and Rcpp frameworks (Eddelbuettel, 2013), GAUSS incorporates a comprehensive 33KG reference panel that encompasses genotype data from over 32,953 individuals across 29 distinct ethnic groups. GAUSS is well equipped to i) estimate ancestral composition, ii) estimate ancestry-informed LD in specified genomic regions, iii) impute summary statistics of unmeasured SNPs, iv) conduct TWAS, and v) correct for the “Winner’s Curse” biases.

To validate its capabilities, we applied GAUSS to the latest PGC Schizophrenia GWAS, leveraging our expansive 33KG panel to estimate ancestry-informed LD. Moreover, we demonstrated that GAUSS can efficiently update even the most recent GWAS data initially imputed using the more limited 1000 Genomes dataset of 2,504 samples. Our 33KG panel refined the dataset and enhanced its resolution significantly.

We believe that, by assisting in accurately analyzing multi-ethnic genetic association studies, GAUSS will help applied researchers in their exploration and understanding of genetic architecture. This could

## Data Availability

All data produced are available online at https://github.com/statsleelab/gauss

https://statsleelab.github.io/gauss/

## GAUSS

potentially lead to the rapid discovery of many novel risk variants/genes contributing to human disease/phenotype.

