## Supplementary Data for "GAUSS: A comprehensive R package for accurate estimation of linkage disequilibrium for variants, Gaussian imputation and TWAS analysis of cosmopolitan cohorts"

Donghyung Lee^1,*^ and Silviu Bacanu

^1^Department of Statistics, Miami University, Oxford, Ohio, USA, ^2^Department of Psychiatry, Virginia Commonwealth University, Richmond, Virginia, USA

**Supplementary Text 1.** The 33KG Reference Panel

GAUSS uses the 32,953 Genomes (33KG) reference panel ^1^. This panel contains 22,691 subjects from the Haplotype Reference Consortium (HRC) ^2^ and 10,262 subjects from CONVERGE ^3^, collectively representing a wide spectrum of ethnic backgrounds: 20,281 Europeans (EUR), 10,800 East Asians (ASN), 522 South Asians (SAS), 817 Africans (AFR), and 533 Native Americans (AMR). Although the HRC is predominantly European, it also incorporates data from 26 diverse populations as part of Phase 3 of the 1000 Genomes Project. Furthermore, an extra EUR population (ORK) was sourced from Orkney Island residents, who are also included in the HRC. In the case of CONVERGE, subjects were grouped into four populations based on their province of origin—China North East (CNE), China Central East (CCE), China South East (CSE) and China Central South (CCS)—all under the broader ASN super-population. A quadratic discriminant analysis model was employed to refine population labels, based on original labels and the first 20 ancestry principal components, potentially reassigning subjects to closely related populations^1^. Overall, the 33KG panel contains 29 populations under five super-populations: AFR, AMR, ASN, EUR, and SAS. Detailed subject counts, abbreviations and population descriptions are provided in Supplementary Table 1.

**Supplementary Table 1.** The population group, the number of subjects, corresponding super population, and description for all 29 population groups in the 33KG reference panel. AFR: African, AMR: Admixed American, ASN: East Asian, EUR: European, SAS: South Asian

| **Population** | **Sample Size** | **Super Population** | **Description** |
| --- | --- | --- | --- |
| ACB | 164 | AFR | African Caribbeans in Barbados |
| ASW | 162 | AFR | African Ancestry in Southwest US |
| BEB | 86 | SAS | Bengali from Bangladesh |
| CCE | 3409 | ASN | China Central East |
| CCS | 2613 | ASN | China Central South |
| CDX | 95 | ASN | Chinese Dai in Xishuangbanna, China |
| CEU | 6360 | EUR | Utah residents with Northern and Western European ancestry |
| CLM | 98 | AMR | Colombians from Medellin, Colombia |
| CNE | 2330 | ASN | China North East |
| CSE | 2020 | ASN | China South-East |
| ESN | 140 | AFR | Esan in Nigeria |
| FIN | 3529 | EUR | Finnish in Finland |
| GBR | 2020 | EUR | British in England and Scotland |
| GIH | 110 | SAS | Gujarati Indian from Houston, Texas |
| GWD | 113 | AFR | Gambian in Western Divisions in the Gambia |
| IBS | 1309 | EUR | Iberian Population in Spain |
| ITU | 95 | SAS | Indian Telugu from the UK |
| JPT | 107 | ASN | Japanese in Tokyo, Japan |
| KHV | 226 | ASN | Kinh in Ho Chi Minh City, Vietnam |
| LWK | 99 | AFR | Luhya in Webuye, Kenya |
| MSL | 87 | AFR | Mende in Sierra Leone |
| MXL | 187 | AMR | Mexican Ancestry from Los Angeles, USA |
| ORK | 5772 | EUR | Orkney Island study |
| PEL | 110 | AMR | Peruvians from Lima, Peru |
| PJL | 121 | SAS | Punjabi from Lahore, Pakistan |
| PUR | 138 | AMR | Puerto Rican in Puerto Rico |
| STU | 110 | SAS | Sri Lankan Tamil from the UK |
| TSI | 1291 | EUR | Toscani in Italia |
| YRI | 52 | AFR | Yoruba in Ibadan, Nigeria |
